# Risk Factors for Hypertension in the Western Highlands of Rural Guatemala

**DOI:** 10.64898/2026.10.14.26365441

**Authors:** Alejandro Chavez, Gabriella Ortiz, Ian Arthur, Sean Duffy

## Abstract

**Objectives:** Uncontrolled hypertension is the leading preventable cause of death and disability worldwide, with nearly two-thirds of affected individuals living in low- and middle-income countries (LMICs). Despite this disproportionate burden, epidemiological data necessary to combat the development of hypertension and its associated complications in LMICs remains scarce.

The objective of this study was to assess risk factors for elevated blood pressure and hypertension diagnosis among adults living in a rural indigenous region of Guatemala.

**Methods:** A cross-sectional analysis was conducted using physician assessment data from 359 patients aged 18 years or older, who participated in a 2022 paired community health worker and physician hypertension study in San Lucas Tolimán, Guatemala. Outcome variables were systolic and diastolic blood pressure, and diagnosed hypertension.

Risk factors examined included age, sex, body-mass index (BMI), smoking status, alcohol use, antihypertensive use, language preference, and other clinical and behavioral characteristics. Multivariable linear regression models were used to assess factors associated with systolic and diastolic blood pressure, while multivariable logistic regression was used for assessing diagnosed hypertension.

**Results:** Higher systolic blood pressure was associated with increasing age, higher BMI, Spanish language preference, and low-intensity physical activity. Diastolic blood pressure was positively associated with increasing BMI, pre-existing diabetes, and Spanish language preference. There was a negative association with high-intensity physical activity. Hypertension diagnosis was associated with increasing age, higher BMI, and pre-existing diabetes.

**Conclusion:** This analysis confirms established hypertension risk factors in a rural indigenous population in Guatemala and identifies additional risk factors that may be relevant in this setting.

## Introduction

Non-communicable diseases (NCDs) account for three-fourths of all deaths globally, with uncontrolled hypertension as the leading preventable cause of death and disability in the world.(1) Uncontrolled hypertension can lead to premature morbidity and life-threatening complications, including stroke, heart failure, and chronic kidney disease.(2) An estimated 1.13 billion people around the world live with hypertension, with two-thirds of these people living in low- and middle-income countries (LMICs).(3,4) Screening and treatment of hypertension has been found to be cost-effective for reducing morbidity and mortality.(5) However, less than 40% of patients with hypertension in LMICs are aware of their condition, and less than 10% of patients have adequate control of their blood pressure.(6) Health systems in LMICs, which face limited access to healthcare, infrastructure, and essential resources, are ill-equipped to address the rising burden of NCDs.

There are several areas of research that could help combat the development of hypertension and other NCDs in LMICs, but significant gaps remain. A key gap is the lack of epidemiological data necessary to determine current prevalence and risk factors in populations, in particular those living in rural and resource-limited areas.(7) Despite the significant burden of disease present in LMICs, funding for research in these countries makes up a small proportion of global research funding.(8) This disparity is evident in countries such as Guatemala, a middle-income country where the burden of NCDs is increasing, ultimately accounting for 59% of deaths in recent reports.(9) Over the past decade, physicians and students affiliated with the University of Wisconsin-Madison have been working with Guatemalan physicians and community health workers (CHWs) to improve capacity for primary care, including chronic disease management and prevention, in San Lucas Tolimán (SLT), a rural municipality in the Western Highlands region of Guatemala. San Lucas has a population of ∼ 30,000 people, of which the majority identify as Kaqchikel Maya.(10) Healthcare services in this region continue to be limited, underscoring the need for local epidemiological data to inform targeted interventions.

A crucial component of our work in Guatemala has been supporting the incorporation of digital health solutions, primarily mobile health (mHealth) applications with integrated clinical decision-support to streamline and expand the healthcare services provided by our CHW partners. In 2020, we created a mobile application using the CommCare platform that enables CHWs serving in the rural communities of San Lucas Tolimán to provide high-quality diagnosis and treatment of hypertension in adults. Importantly, these digital platforms facilitate further epidemiological analysis and ongoing program monitoring, helping address the lack of data on hypertension and its associated risk factors.(11) As part of a larger initiative to improve chronic disease management in the region, a hypertension diagnostic study was conducted in 2022, in which CHWs and physicians conducted paired assessments of 359 patients and reached independent diagnoses regarding hypertension status.(12) We utilized the data collected from the physician portion of these paired assessments to investigate the growing burden of hypertension in this population. Accordingly, the objective of this study was to assess prevalent risk factors for an elevated blood pressure and hypertension diagnosis among adults living in the rural indigenous communities of San Lucas Tolimán, Guatemala.

## Materials and Methods

### Data Collection

For this cross-sectional study, we used de-identified data collected as part of a non-inferiority study with IRB approval via the University of Wisconsin Department of Family Medicine and Community Health (IRB protocol number 2021-1462, approved 11/30/21). The aim of the parent trial (“Assessing the accuracy of community health workers for hypertension diagnosis”) was to demonstrate that the test intervention (ie, community health worker-led diagnosis of hypertension facilitated by a mobile application with decision support) is non-inferior to the comparator and standard of care (ie, physician-based diagnosis).(12) In the parent trial, interested participants were initially screened for eligibility, consented if eligible and in agreement to participate, and then enrolled. Data was collected using tablet devices running a CommCare application. Data collected included sex, date of birth, community, language preference, self-reported medical history, use of antihypertensives, physical activity, type of stove used at home and its location, smoking and alcohol use history, blood pressure, pulse, weight, and height. The application would then suggest a determination of the patient’s hypertension status (yes, no, or possible with followup needed) and the user (CHW or physician) could choose to agree or disagree and if they disagree, would then provide their own determination. For patients with possible hypertension, a single follow-up visit was indicated to determine hypertension status, during which blood pressure and weight measurements were repeated.

### Patient Demographics

The study population was non-pregnant individuals who were 18 years or older who consented to participate in the study. The participants all lived in the rural communities of San Lucas Tolimán, Sololá, Guatemala, a predominantly indigenous Mayan population and spoke either Spanish or Kaqchikel. See table 2 in the Results section for characteristics of the study sample. They were recruited to participate in the non-inferiority study by the CHWs who live in the communities they serve and helped conduct the study visits. Study visits occurred at central locations within the target communities. This analysis uses demographic data collected by a CHW during participant registration and clinical data collected by a physician during the aforementioned non-inferiority study. Clinical data collected from the CHW assessments were not used.

### Specific Protocols

Determination of hypertension diagnosis by the application was based on well-established criteria for hypertension diagnosis from the World Health Organization (WHO) and the International Society of Hypertension.(13),(14) In the absence of a previous diagnosis of hypertension and the patient not currently taking an antihypertensive medication, a blood pressure of 140/90 or greater required a follow-up visit with another measurement of 140/90 or greater to confirm the diagnosis. Two elevated measurements at distinct visits were also used to confirm diagnosis of a patient with a previous diagnosis of hypertension who was not currently taking an antihypertensive medication, as well as for a patient without a previous diagnosis, but who was currently taking an antihypertensive medication. The presence of a previous diagnosis of hypertension and the patient currently taking an antihypertensive medication was sufficient to confirm a diagnosis of hypertension regardless of blood pressure. A blood pressure of 180/110 or greater was sufficient to confirm a diagnosis of hypertension during a single visit regardless of previous diagnosis and whether the patient is taking an antihypertensive medication.

Blood pressure measurements were taken with an Omron automated blood pressure measurement device that was validated by the STRIDE BP initiative with patients sitting for at least 5 minutes before initial measurement, and additional measurements were taken, if elevated, per International Society of Hypertension (ISH) guidelines.(15) If the initial measurement was above or equal to 130/85, two additional measurements were taken with a minimum of one minute between subsequent measurements and the mean of the final two measurements was used as the recorded blood pressure for each participant.

### Overview of considered hypertension risk factors

The outcomes (response variables) we included in our analyses were systolic and diastolic blood pressures, as well as hypertension diagnostic status. Risk factors (explanatory variables) of interest included patient sex, age, body-mass index (BMI), current smoking status, recent alcohol use, preferred language, physical activity measures, the type of stove for cooking and its location in the home, and history of kidney disease, diabetes, and hyperlipidemia. We included the covariates of recent use of antihypertensive medication and prior diagnosis of hypertension to control for their impact on systolic and diastolic blood pressure. Further details of the considered risk factors can be found in Table 1.

**Table 1.** Overview of considered risk factors for multivariable regression analysis, San Lucas Tolimán, Guatemala, 2022. Source: Table created by authors.

| Variable | Type | Details |
| --- | --- | --- |
| Current smoking status | Categorical: yes, no | Answer to the question: “Do you smoke?” |
| Recent alcohol use | Categorical: yes, no | Refers to the past 30 days. Data was captured quantifying alcohol use, but given overall minimal alcohol use and low amounts when present, the binary question was used instead. |
| Recent use of antihypertensive medication | Categorical: yes, no | Refers to the past 15 days. |
| Pre-existing diagnoses (hypertension, kidney disease, diabetes, hyperlipidemia) | Categorical: yes, no | Self-reported by patient and were not confirmed with any lab testing or reviewing of other medical records. |
| Patient Sex | Categorical: male, female |  |
| Preferred language | Categorical: Spanish, Kaqchikel | Kaqchikel is the predominant indigenous Mayan language in the region. |
| Type of stove used for cooking and location | Categorical: Concatenation of two separate fields with one referring to the type of stove (gas, wood burning without chimney, and wood burning with chimney) and the other its location (indoor in main living area, indoor but separate from main living area, and outdoor) | Given this field is meant to be a proxy for smoke exposure from cooking, the two fields must be looked at in conjunction. |
| Low-intensity exercise | Continuous/Numerical: 0-7 | Number of days over previous week that participant did at least 30 minutes of normal walking, climbing hills/stairs, collecting/carrying firewood, or working in the garden/fields |
| High-intensity exercise | Continuous/Numerical: 0-7 | Number of days over previous week that participant did at least 30 minutes of brisk walking, running, biking, or playing soccer |
| Patient Age | Continuous/Numerical | Calculated from date of birth if known, otherwise from patient reported age |
| Patient BMI | Continuous/Numerical | Calculated from measured weight and height during visit |

**Table 2.** Overview of study participant demographic including mean and standard deviation, San Lucas Tolimán, Guatemala, 2022. Source: Findings from parent study with table created by authors.

| Variable | Outcome<br>Mean and SD where applicable |
| --- | --- |
| Total participants, n | 359 |
| Diagnosed hypertension, n (%) | 55 (15% of total) |
| Systolic blood pressure (mmHg) | 116 ± 18 (range: 80-219) |
| Diastolic Blood Pressure (mmHg) | 73.8 ± 9.8 (range: 52-113) |
| Pre-existing diabetes, n (%) | 43 (12% of total) |
| Age (years) | 46 ± 16 (range: 18-86) |
| BMI (kg/m<sup>2</sup>) | 29.2 ± 5.5 (range: 15.1-58.7) |
| Sex (Male, Female), n (%) | M 61 (17% of total), F 298 (83% of total) |
| Language (Spanish, Kaqchikel), n (%) | S 160 (45%), K 199 (55%) |
| Low-intensity exercise | 4.42 ± 3.07 (range 0-7) |
| High-intensity exercise | 0.33 ± 1.19 (range 0-7) |
| Medical history |  |
| Hyperlipidemia, n (%) | 30 (8% of total) |
| Kidney disease, n (%) | 9 (3% of total) |
| Smoking status, n (%) | 11 (3% of total) |
| Alcohol use | 27 (8% of total) |
| Stove usage |  |
| Inside with gas, n (%) | 43 (12% of total) |
| Inside with wood, no chimney, n (%) | 7 (2% of total) |
| Inside with wood, chimney, n (%) | 234 (65% of total) |
| Separate room with gas, n (%) | 2 (0.5% of total) |
| Separate room with wood, no chimney, n (%) | 4 (1% of total) |
| Separate room with wood, chimney, n (%) | 52 (15% of total) |
| Outside with wood, no chimney, n (%) | 1 (0.3% of total) |
| Outside with wood, chimney, n (%) | 16 (5% of total) |
| Recent use of antihypertensive, n (%) | 28 (7.8% of total) |

### Statistical approach for hypertension risk factors

Multivariable linear regression models were used to analyze the relationship between the outcome variables of systolic and diastolic blood pressure with the risk factors detailed in the previous section. Of note, if a patient underwent two visits in the parent study, the blood pressure from the second visit was used.

A multivariable logistic regression model was used to analyze the relationship between the outcome variable of hypertension diagnostic status (“yes” or “no”) with the risk factors detailed in the previous section. Pre-existing diagnosis of hypertension was not included as a possible risk factor in this analysis given that it was a key component of the diagnostic determination for hypertension. The hypertension diagnostic status was determined by the physician during the initial visit or during a follow-up visit if deemed necessary by the physician.

R was used for statistical analysis, for which the base “stats” package was used to determine the optimal regression models. For linear regression, the “lm” function was first used and for logistic regression the “glm” function was first used. The “step” function was then used to identify the optimal model based on the Akaike Information Criterion (AIC) score.(16) The “step” function iteratively adds or removes predictor variables to find the best fitting model, which is identified by having the lowest AIC score.

## Results

### Analysis of risk factors for elevated systolic blood pressure

The optimal multivariable linear regression model with systolic blood pressure as the outcome included the following variables all showing a positive correlation: increasing BMI, self-reported diagnosis of diabetes, preferred language of Spanish, increasing age, increasing amount of low-intensity exercise, self-reported diagnosis of hypertension, and current use of an antihypertensive medication. Details of this multivariable regression analysis can be found in Table 3. This optimal model had an adjusted R-squared value of 0.345. All variables included in the model, with the exception of preferred language, were shown to be statistically significant with a p-value less than 0.05.

**Table 3.**
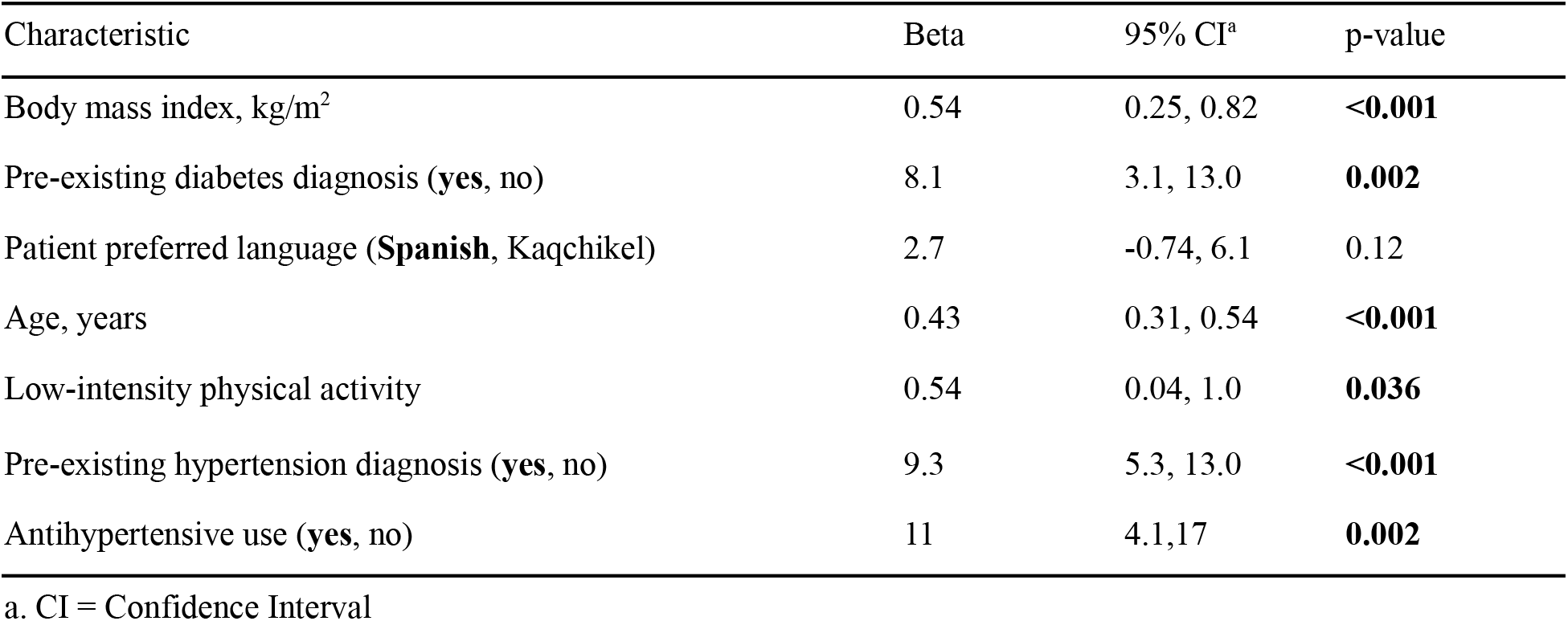
Multivariable linear regression analysis of risk factors associated with elevated systolic blood pressure, San Lucas Tolimán, Guatemala, 2022. Source: Findings from parent study with table created by authors.

### Analysis of risk factors for elevated diastolic blood pressure

The optimal multivariable linear regression model with diastolic blood pressure as the outcome included the following variables all showing a positive correlation: increasing BMI, self-reported diagnosis of diabetes, preferred language of Spanish, increasing age, self-reported diagnosis of hypertension, and current use of an antihypertensive medication. The optimal model also included a variable with negative correlation: increasing amount of high-intensity exercise. Details of this multivariable regression analysis can be found in Table 4. This optimal model had an adjusted R-squared value of 0.2193. All variables included in the model, with the exception of preferred language and amount of high-intensity exercise, were shown to be statistically significant with a p-value less than 0.05.

**Table 4.** Multivariable linear regression analysis of risk factors associated with elevated diastolic blood pressure, San Lucas Tolimán, Guatemala. Source: Findings from parent study with table created by authors.

| Characteristic | Beta | 95% CI<sup>a</sup> | p-value |
| --- | --- | --- | --- |
| Body mass index, kg/m<sup>2</sup> | 0.43 | 0.26, 0.60 | **<0.001** |
| Pre-existing diabetes diagnosis (**yes**, no) | 6.0 | 3.1, 8.9 | **<0.001** |
| Patient preferred language (**Spanish**, Kaqchikel) | 1.40 | -0.51, 3.3 | 0.20 |
| High-intensity physical activity | -0.76 | -1.5, 0.03 | 0.058 |
| Pre-existing hypertension diagnosis (**yes**, no) | 4.60 | 2.2, 6.9 | **<0.001** |
| Antihypertensive use (**yes**, no) | 6.2 | 2.4, 10 | **0.001** |

a. CI = Confidence Interval

### Analysis of risk factors for hypertension diagnosis

The optimal multivariable logistic regression model with hypertension diagnosis as the outcome included the following variables all showing a positive correlation: increasing BMI, self-reported diagnosis of diabetes, increasing age, and self-reported diagnosis of hyperlipidemia. This optimal model had an AIC score of 273.28. Age and self-reported diagnosis of diabetes were shown to be statistically significant predictors with a p-value less than 0.05, while BMI and self-reported diagnosis of hyperlipidemia were not statistically significant. Details of this multivariable regression analysis can be found in Table 5.

**Table 5.** Multivariable logistic regression analysis of risk factors associated with hypertension diagnosis, San Lucas Tolimán, Guatemala, 2025. Source: Findings from parent study with table created by authors.

| Characteristic | OR<sup>a</sup> | 95% CI<sup>b</sup> | p-value |
| --- | --- | --- | --- |
| Body mass index, kg/m<sup>2</sup> | 1.05 | 0.99, 1.11 | 0.10 |
| Pre-existing diabetes diagnosis (**yes**, no) | 3.43 | 1.62, 7.18 | **0.001** |
| Age, years | 1.05 | 1.03,1.07 | **<0.001** |
| Pre-existing hyperlipidemia diagnosis (**yes**, no) | 2.13 | 0.81, 5.27 | 0.11 |

a. OR = odds ratio
b. CI = confidence interval

## Discussion

In this cross-sectional analysis of adults living in the rural Indigenous communities of San Lucas Tolimán, Guatemala, we identified several factors associated with elevated blood pressure and a diagnosis of hypertension. Increasing age, higher BMI, and self-reported diabetes were consistently associated with higher systolic and diastolic blood pressure as well as with a hypertension diagnosis. These findings align with established hypertension risk factors observed globally and reinforce the growing burden of cardiometabolic disease given lower quality diets, increasingly sedentary lifestyles, and aging populations across the world. (6,17,18) The effects of this growing disease burden are most consequential in LMICs where limitations of the healthcare system prevent an adequate response to manage these conditions and their complications for the most impacted and vulnerable members of their populations. Importantly, this study contributes locally derived epidemiologic data from an underserved Indigenous population for whom such data remain scarce.

A recent analysis done by our team published in 2024 using electronic health record (EHR) data found a prevalence of hypertension of 16.7% within this population, which was in line with previous estimates in the nearby region and comparable to our finding of 15% prevalence.(19) The previous analysis also investigated risk factors using the EHR data and found similar findings to those in this study. There was a positive correlation between male sex, increasing age, and increasing weight with systolic blood pressure. Increasing age and increasing BMI were found to have significant positive correlation with systolic blood pressure in this study as well. When using diastolic blood pressure as the outcome, history of diabetes and increasing weight were positively correlated. These same findings were seen in this study with BMI used instead of weight. These findings are consistent with other studies conducted in similar low- and middle-income settings, which have identified age, adiposity, and metabolic comorbidities such as diabetes as key risk factors for hypertension.(20),(21) This analysis re-evaluates these risk factors as well as others with standardized, cross-sectional data collection.(19)

### Key Findings and Interpretation

Pre-existing history of diabetes emerged as a strong predictor with high levels of statistical significance and large effect sizes across both blood pressure and diagnosis of hypertension. This highlights the need for dual-screening of both conditions and makes an argument for integrated chronic disease management given the significant overlap in complications for both conditions, including acute coronary syndrome, stroke, peripheral arterial disease, and chronic kidney disease. This association is likely driven by the shared pathophysiology mechanisms, including insulin resistance, endothelial dysfunction, and activation of the renin-angiotensin-aldosterone system.(22) Collectively, the coexistence of these conditions substantially increases cardiovascular risk.(23),(24)

Increasing patient age was a strong predictor for systolic blood pressure and a diagnosis of hypertension. Increasing BMI emerged as a strong predictor for both systolic and diastolic blood pressures. These findings showcase the contribution of aging and increasing adiposity to the development of hypertension. A recent joint report from the American College of Cardiology (ACC) and American Heart Association (AHA) notes that there is an association between blood pressure with “fatal and nonfatal cardiovascular events in a graded, log-linear fashion, with an approximate doubling of risk for each 20-mm Hg higher SBP and 10-mm Hg higher DBP level.”(17) This highlights the need to both prevent the development of hypertension, but also lower blood pressure for those with an established hypertension diagnosis, especially the most uncontrolled, given the increased risk of cardiovascular complications, as well as other established complications, including chronic kidney disease and stroke. While age is non-modifiable, it speaks to the need to screen in this population and targeted interventions given the compounding risk for the elderly with uncontrolled hypertension, especially for those with an elevated BMI.

Given hyperlipidemia was included in the optimal logistic regression model for hypertension diagnosis, but not in the optimal linear regression models for blood pressure, it may indicate increased healthcare contact for those particular patients with hyperlipidemia allowing for a diagnosis of hypertension instead of an actual physiological relationship between hyperlipidemia and blood pressure. In addition, given that medical history used in this analysis was self-reported without any lab-based verification, it is likely there is underreporting of true rates of hyperlipidemia, as well as other comorbidities.

### Physical Activity and Behavioral Factors

There was a mixed relationship between physical activity and blood pressure outcomes. Of note, physical activity measures were defined as the number of days over the past week the participant completed at least 30 minutes of the given physical activity. For systolic blood pressure there was a positive correlation with low-intensity physical activity, meaning the more low-intensity physical activity a participant did, the higher their systolic blood pressure. For diastolic blood pressure there was a negative correlation with high-intensity physical activity, meaning the more high-intensity physical activity a participant did, the lower their diastolic blood pressure. These physical activity measures are self-reported and participants may have overestimated their amount of low-intensity physical activity vs high-intensity physical activity. The positive correlation in this study between systolic blood pressure and low-intensity physical activity is inconsistent with the established relationship of decreasing blood pressure with increasing levels of physical activity. A meta-analysis showed that aerobic exercise reduced blood pressure by 3/2.4 mmHg and that the effect was independent of any weight loss.(25) Confounding by indication is likely at play here given those with a diagnosis of hypertension, especially those whose blood pressure is most elevated, are most likely to have been recommended to increase physical activity by a medical provider to improve control of their hypertension. In addition we are only capturing a blood pressure measurement at a single point of time instead of looking at trends over time.

### Language Preference and Social Context

A patient’s preferred language was meant to serve as a proxy for healthcare access due to possible language barriers as well as general socioeconomic factors. The options were either Spanish or the predominant Mayan indigenous language, Kaqchikel. While not statistically significant, Spanish language was included in the optimal linear regression model with a positive correlation for both systolic and diastolic blood pressures. The medical system in Guatemala is generally Spanish-dominated with Spanish as the predominant language for medical providers as well as patient-facing resources. It may be that Spanish language is a marker of greater acculturation to a Western diet that aligns with an elevated BMI and as seen in this study, a positive correlation with elevated blood pressure. The question was a single-select multiple choice asking for preferred language making it unable to differentiate between monolingual and bilingual participants. Future studies that adequately capture additional measures of socioeconomic status, such as level of education, type of occupation, and household income may provide insight into possible relationships to blood pressure.

### Limitations

Limitations of this study include that several key variables were self-reported by the patient, including pre-existing diagnoses, introducing the potential for misclassification. A thorough medication history and laboratory verification would have provided more confidence in these data. Factors related to smoke exposure via stoves may not adequately capture previous exposures given the data only reflects the current exposure. Of note, there have been several initiatives to transition wood-burning stoves without chimneys to wood-burning stoves with chimneys over the past 10 to 20 years in the region. In addition, this study only included data of those patients that participated in the non-inferiority study, which may not fully represent the patient population as a whole. For example, of the 359 participants, only 61 were male likely due to unavailability given the timing of the visits were during typical hours of agricultural work, which is primarily done by males. Selection bias may have also been introduced due to the possibility of having participants who were most concerned about having hypertension or being previously told they had an elevated blood pressure. A future study could overcome a possible selection bias by randomizing subject selection across a geographical area.

## Conclusion

In a rural Indigenous population in the Western Highlands of Guatemala, elevated blood pressure and hypertension were strongly associated with increasing age, elevated BMI, and diabetes. These findings highlight the urgent need for locally informed and integrated chronic care management, in particular for hypertension and diabetes, in similar types of settings across LMICs. CHW-led care models may be in the best position to address existing clinical care gaps, and also contribute to severely lacking epidemiological data needed to develop targeted interventions. Future research can delve further into how socioeconomic factors play a role in the development of hypertension and other NCDs in order to inform the development of impactful models for chronic disease management.

## Data Availability

All data produced in the present study are available upon reasonable request to the authors.

## Statements and Declarations

Research reported in this publication was supported by the Fogarty International Centre of the National Institutes of Health under award number R21TW011891. The content is solely the responsibility of the authors and does not necessarily represent the official views of the National Institutes of Health. The NIH was not involved in study design or execution, nor in the decision to submit for publication.

## Conflict of interest: the authors declare no potential conflict of interest.

All authors contributed to the study conception and design. Material preparation, data collection and analysis were performed by Alejandro Chavez and Gabriella Ortiz. Supervision, interpretation of findings, and critical review of the manuscript was performed by Sean Duffy. Critical review, feedback, and revision of the manuscript was performed by Ian Arthur. The first draft of the manuscript was written by Alejandro Chavez and Gabriella and all authors commented on previous versions of the manuscript. All authors have reviewed and approved the final manuscript and agree to be accountable for the work and its integrity.

## Ethics approval and consent to participate

research was performed in accordance with the Declaration of Helsinki. This project was reviewed and approved by the San Lucas Healthcare Committee, as well as the University of Wisconsin IRB (submission ID 2022-0794).

Informed consent was obtained from all individual participants included in the study.

## Consent for publication

not applicable.

